# Chronic Traumatic Encephalopathy Neuropathologic Change is Infrequent Following Isolated Moderate-Severe Traumatic Brain Injury

**DOI:** 10.1101/2025.02.24.25322800

**Authors:** E. Selmanovic, A. Pruyser, A.C. Seifert, B.N. Delman, E.L. Thorn, R. Folkerth, K. Dams-O’Connor

**Affiliations:** Friedman Brain Institute, Icahn School of Medicine at Mount Sinai, New York, NY; Department of Rehabilitation and Human Performance, Icahn School of Medicine at Mount Sinai, New York, NY; Nash Family Department of Neuroscience, and Artificial Intelligence & Human Health, Ronald M. Loeb Center for Alzheimer’s Disease, Friedman Brain Institute, Icahn School of Medicine at Mount Sinai, New York, NY; Department of Diagnostic, Molecular and Interventional Radiology, Biomedical Engineering and Imaging Institute, Graduate School of Biomedical Sciences, Icahn School of Medicine at Mount Sinai, New York City, NY; Department of Diagnostic, Molecular and Interventional Radiology, Icahn School of Medicine at Mount Sinai, New York City, NY; Department of Pathology, Molecular, and Cell Based Medicine, Ronald M. Loeb Center for Alzheimer’s Disease, Friedman Brain Institute, Icahn School of Medicine at Mount Sinai, New York, NY; Neuropathology Brain Bank & Research CoRE, Mount Sinai Hospital, New York, NY; Department of Neurology, Icahn School of Medicine at Mount Sinai, New York, NY

**Keywords:** Autopsy - Brain trauma –Neurodegeneration – Tauopathy – Chronic Traumatic Encephalopathy

## Abstract

Chronic traumatic encephalopathy-neuropathologic change (CTE-NC) has been primarily studied in contact sport athletes with repetitive head impacts (RHI). An association with isolated traumatic brain injury (TBI) is less clear. We systematically reviewed the autopsied cohort of Late Effects of TBI (LETBI), characterized primarily by isolated TBI but also including RHI, for features of CTE-NC. A consecutive series of 44 brains underwent comprehensive neuropathologic evaluation, exceeding recommended CTE consensus protocols. Of the 44, 6 (13.6%) (age range, 3^rd^-7^th^ decades; median, 6^th^ decade) had CTE-NC, forming the basis for this exploratory analysis. Ex vivo neuroimaging (in 4 of 6) highlighted traumatic and white matter microvascular lesions, facilitating histological sampling of subtle neuropathologies that might otherwise have been missed by conventional sectioning. Macroscopically, 5 had cortical (contusional) and white matter (torsional) volume loss, with septal lesions and hydrocephalus ex vacuo (i.e., structural lesions of moderate-severe TBI). Microscopically, tau-immunopositive neuronal and astrocytic pathology in a perivascular arrangement within sulcal depths met current pathognomonic criteria for CTE-NC in 5 (3 “low” and 2 “high” burden); 1 had more limited findings considered “suspicious” for CTE-NC. Five of 6 cases with any CTE-NC reported substantial exposure to RHI, through contact sport ranging over at least 16 years. One case had no known exposure to RHI: this case (death: 6th decade) had 2 isolated severe TBIs (sustained 30y and 3y prior to death). Of note, one case with “high” CTE also had Alzheimer Disease Neuropathologic Change (high stage), Lewy Body Disease (limbic), and TDP43 accumulation (“polyproteinopathy”). Aging-related tau astrogliopathy, mostly subpial, was seen in 4 cases. Glial tau was also noted around old cavitary contusions in 2. These findings converge with prior studies demonstrating that CTE is largely associated with RHI and is infrequent among individuals with isolated TBI.

## INTRODUCTION

Chronic traumatic encephalopathy (CTE) is a constellation of neurodegenerative pathology associated with head trauma, primarily identified in contact sport athletes with extensive repetitive head impacts (RHI). Originally described in boxers as “dementia puglistica”,^1^ the term CTE now refers to a specific pattern of pathology that has been detected in decedents who engaged in a wide variety of contact sports,^2–7^ including before or while engaged in military training or combat (though not to date from military exposure per se),^2,8–10^ behavior-related etiologies of RHI such as head banging,^2,11,12^ and, in a singular case report, intimate partner violence (IPV).^13^

In 2015, a consensus panel of neuropathologists defined CTE neuropathologic change (CTE-NC) by the “accumulation of abnormal hyperphosphorylated tau (p-tau) in neurons and astroglia distributed around small blood vessels at the depths of cortical sulci and in an irregular pattern”.^14^ A second meeting of the panel later specified that “the perivascular p-tau aggregates should include neurofibrillary tangles, with or without astrocytes, and that the focus had to be in deeper cortical layers not restricted to subpial and superficial regions”; this refinement was to preclude the erroneous interpretation of “aging-related tau astrogliopathy” (ARTAG) as CTE-NC.^15^

Both consensus papers identified supplementary supportive, but not independently diagnostic, features including preferential involvement of hippocampal CA2 and CA4 regions, contrasting with the usual pattern of CA1 and subicular involvement in Alzheimer Disease (AD), as well as p-tau foci in subcortical nuclei.^14,15^ Macroscopic septal abnormalities, and transactive response DNA-binding protein 43 (TDP-43) pathology, affecting the hippocampus, anteromedial temporal cortex, and amygdala were also delineated as supportive features.^14,15^

While four presumably “progressive” stages of pathological severity were first described in 2013,^2^ the suspected progression has not yet been validated independently and, instead, the second consensus proposed a severity grade that characterizes cases as “low” or “high” CTE-NC burden, based upon examination of neuronal p-tau using specific neuroanatomic sampling and tissue staining.^15^

In the years since consensus criteria were established, CTE-NC has been reported in as many as 87% of former contact sport athletes recruited specifically for exposure to RHI.^16^ However, studies analyzing brain bank cohorts have reported low levels of CTE-NC (12-31.8%), nearly always in individuals with history of contact sports,^3,17^ and only rarely seen in cases without known RHI exposure.^18–20^ Additionally, while old and/or recent TBI was identified grossly or microscopically in all 14 women with IPV in a prospectively accrued cohort, no CTE-NC was detected on comprehensive neuropathologic analysis.^21^ Given that these populations are not always representative of the general population, the actual prevalence of CTE-NC in unselected samples requires further investigation. In particular, few studies have investigated the long-term sequelae of non-repetitive (i.e., isolated or single) head trauma, ranging from mild to severe exposures, specifically as it pertains to CTE. The minimum threshold and mechanism of head trauma exposure required to precipitate CTE-NC is unclear – and it is not known whether a single TBI can initiate CTE-NC.^22^ The long-term clinical and neuropathological sequelae of isolated TBI are of growing public health interest,^23–26^ and in-vivo biomarker discovery efforts to date have defined “chronic TBI” as referring to RHI or isolated TBI as though they are interchangeable.^27^ It is essential to determine whether the neuropathological signatures of RHI and TBI differ, particularly in light of evidence suggesting their clinical sequelae may be similar.^28^

We sought to determine the prevalence of CTE-NC and any co-occurring neuropathology following a spectrum of head trauma exposures in the Late Effects of TBI (LETBI) brain donor cohort, all of whom are subjected to a comprehensive diagnostic protocol of brain sampling at autopsy.^29^ A consecutive case series of 44 brains with a range of head trauma exposure patterns including RHI, isolated TBI or a combination of etiologies were extensively examined for the presence of CTE and other neurodegenerative pathology. We provide evidence that CTE-NC is infrequent in cases of isolated TBI, implying that it instead may be largely related to RHI.

## METHODS

### Case recruitment and clinical evaluation

A consecutive series of 44 autopsied cases were collected for the LETBI study at the Icahn School of Medicine at Mount Sinai, as previously described.^21,29^ Brain donors are individuals who enroll in the LETBI study during life^29^ therein making known their wishes for brain donation; individuals whose families contact the study team to participate in brain donation at the time of death; or decedents with a history of TBI identified by local medical examiners and referred to our center. All cases underwent comprehensive medical and public record review (i.e., hospital documentation, obituaries, yearbooks, etc.) to determine TBI/RHI history and study eligibility; additional retrospective informant interviews were completed on 28 cases. All records and data were reviewed (by KDOC, AP, ES, and RDF) to confirm head/brain trauma exposure characterization and medical, psychological, and other comorbidities. Study procedures were approved and deemed not human subjects research (NHSR) by the Mount Sinai Program for the Protection of Human Subjects.

### Neuropathologic evaluation

All specimens were fixed in 10% formalin for a minimum of 4 weeks prior to the completion of a multimodal neuropathological evaluation, as detailed previously.^21,29,30^ Thirty-six cases underwent ex vivo 3 Tesla magnetic resonance imaging (MRI) to assist in the identification of subtle or small lesions for diagnostic microscopic evaluation.^21,29^ Each imaging dataset was reviewed by a board-certified CAQ neuroradiologist (BND) with expertise in the interpretation of ex vivo MRI. Findings of potential interest were marked for correlative histologic sampling.

In all cases, histologic sampling followed standardized protocols for evaluation of neurodegenerative disease, including CTE-NC, with histochemical and immunohistochemical (IHC) staining.^14,15^ Briefly, for each case, comprehensive sectioning exceeded all anatomical regions required per CTE consensus criteria (blocks submitted, 23-65 per case, median 42). Sections from these regions were then prepared for standard Luxol-fast blue/hematoxylin and eosin staining (LHE) and for IHC employing antibodies to p-tau (AT8, Invitrogen, catalogue # ENMN1020, concentration 1:1000), amyloid-beta (6E10, BioLegend, catalogue # 803015 concentration 1:5000), phospho-TDP (1D3): BioLegend, catalogue # 829901 concentration 1:5000), and alpha-synuclein (MilliporeSigma, catalogue # MABN824MI, concentration 1:3000).

Additional sections were prepared for Perls’ Prussian blue stain for hemosiderin using standard protocols. Immunostains for p62 (BD Biosciences, catalogue #610832, concentration 1:250) were performed per the manufacturer’s directions.

Alzheimer disease-neuropathologic change (AD-NC) was characterized using the “ABC” system,^31^ recognizing that the amyloid and tau depositions did not always match the recognized patterns typically seen in cases selected for dementia (e.g., those studied in AD brain banks). This system provided a semi-quantitative method for summarizing tau (neuronal in any amount), amyloid, and plaque burden to facilitate comparison among cases. Aging-related tau astrogliopathy (ARTAG) was evaluated per published guidelines.^32^

### Statistical analysis

No statistical analysis plan was utilized for this hypothesis-generating diagnostic case series. All demographic characteristics, relevant clinical histories and neuropathological findings are provided in a descriptive format.

## RESULTS

### Demographic and clinical information

Forty-four cases were collected prospectively from 2018 to 2024 (Table 1). Ages ranged from 2nd to 10^th^ decade with a median age of 54 (exact ages not provided to protect privacy). Thirty-four (77%) cases were male and twenty-four (54%) were white; 8 (18%) were of Hispanic ancestry. Medical, family interview and self-report records indicate that 17 (38.6%) cases had a history of RHI only (7 from contact sports only, 4 from military service only, 4 from IPV only, 1 from contact sport and self-harm, and 1 from military service and IPV). 13 (29.5%) had isolated TBI only, and 15 (34%) had mixed head trauma exposure (i.e., RHI and isolated TBI).

**Table 1.**
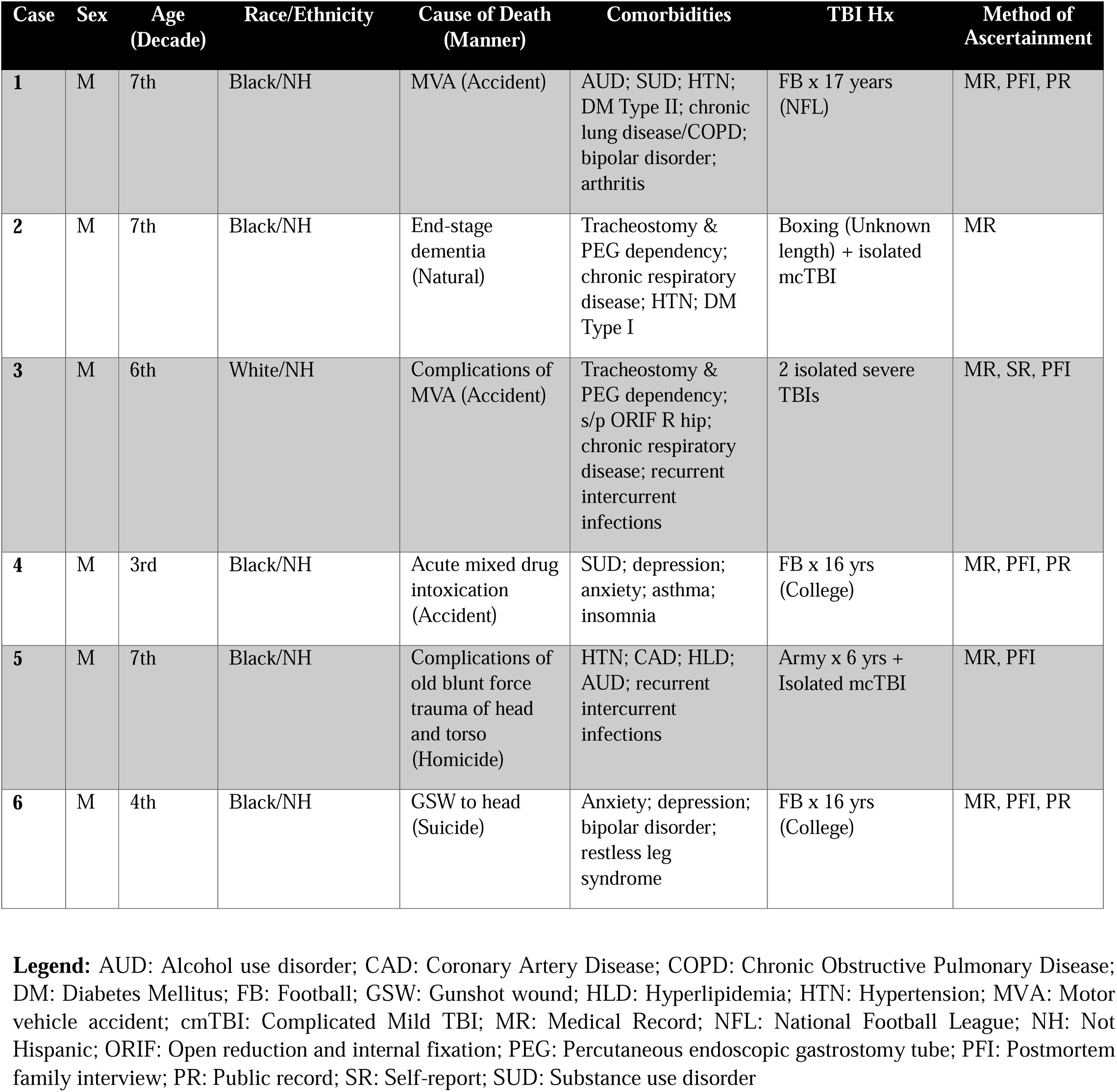
Clinical history for cases with CTE-NC in the LETBI cohort.

| Case | Sex | Age<br>(Decade) | Race/Ethnicity | Cause of Death<br>(Manner) | Comorbidities | TBI Hx | Method of<br>Ascertainment |
| --- | --- | --- | --- | --- | --- | --- | --- |
| 1 | M | 7th | Black/NH | MVA (Accident) | AUD; SUD; HTN;<br>DM Type II; chronic<br>lung disease/COPD;<br>bipolar disorder;<br>arthritis | FB x 17 years<br>(NFL) | MR, PFI, PR |
| 2 | M | 7th | Black/NH | End-stage<br>dementia<br>(Natural) | Tracheostomy &<br>PEG dependency;<br>chronic respiratory<br>disease; HTN; DM<br>Type I | Boxing (Unknown<br>length) + isolated<br>mcTBI | MR |
| 3 | M | 6th | White/NH | Complications of<br>MVA (Accident) | Tracheostomy &<br>PEG dependency;<br>s/p ORIF R hip;<br>chronic respiratory<br>disease; recurrent<br>intercurrent<br>infections | 2 isolated severe<br>TBIs | MR, SR, PFI |
| 4 | M | 3rd | Black/NH | Acute mixed drug<br>intoxication<br>(Accident) | SUD; depression;<br>anxiety; asthma;<br>insomnia | FB x 16 yrs<br>(College) | MR, PFI, PR |
| 5 | M | 7th | Black/NH | Complications of<br>old blunt force<br>trauma of head<br>and torso<br>(Homicide) | HTN; CAD; HLD;<br>AUD; recurrent<br>intercurrent<br>infections | Army x 6 yrs +<br>Isolated mcTBI | MR, PFI |
| 6 | M | 4th | Black/NH | GSW to head<br>(Suicide) | Anxiety; depression;<br>bipolar disorder;<br>restless leg<br>syndrome | FB x 16 yrs<br>(College) | MR, PFI, PR |

**Legend:** AUD: Alcohol use disorder; CAD: Coronary Artery Disease; COPD: Chronic Obstructive Pulmonary Disease; DM: Diabetes Mellitus; FB: Football; GSW: Gunshot wound; HLD: Hyperlipidemia; HTN: Hypertension; MVA: Motor vehicle accident; cmTBI: Complicated Mild TBI; MR: Medical Record; NFL: National Football League; NH: Not Hispanic; ORIF: Open reduction and internal fixation; PEG: Percutaneous endoscopic gastrostomy tube; PFI: Postmortem family interview; PR: Public record; SR: Self-report; SUD: Substance use disorder

Of the 44 cases, 6 (13.6%) were found to have tau-immunopositive neuronal and astrocytic pathology located in a perivascular arrangement in sulcal depths, with 5 meeting current criteria for CTE-NC, and 1 with very limited findings considered “suspicious” for CTE-NC (Case 4) (Table 2; Figure 2). The ages of this subgroup ranged from the 3rd to 8th decade with a median age of 6^th^ decade. Of the 5 cases considered diagnostic, 3 were rated as “low” CTE-NC (Cases 3, 5, and 6) and 2 as “high” CTE-NC (Cases 1 and 2) based on the distribution of neuronal tau throughout cortical and subcortical regions (Table 2; Figure 2) (see below for additional neuropathologic characteristics of these cases). CTE-NC was not found in any of the remaining 38 cases, which will not be further discussed in this report.

**Table 2.** Postmortem MRI, macroscopic and histologic findings for cases with CTE-NC in LETBI cohort.

| Case | Brain weight (g)* | Ex vivo imaging | Old TBI pathology | CTE-NC | AD-NC | ARTAG | Atherosclerosis | Arteriolosclerosis | Other |
| --- | --- | --- | --- | --- | --- | --- | --- | --- | --- |
| 1 | 1001 | - | - | High | A0B1C0 (PART) | + (subpial) | Moderate | Moderate | Limited gross examination (handling artifact) |
| 2 | 1150 | + (periventricular WM hyperintensity; torn septum; bilateral parieto-occipital cortical hyperintensity) | SDH; torn septum, WM atrophy with hydrocephalus ex vacuo (TAI) | High | A3B3C2 | + (subpial) | - | Mild | FTLD with TDP-43 with p62; LBD (limbic); old watershed ischemia |
| 3 | 1307 | + (hemosiderin and volume loss, frontotemporal; linear WM defects, SFG, with WM hyperintensities; torn septum; ventricular expansion) | Contusions; HTTs, torn septum and thin CC, WM atrophy with hydrocephalus ex vacuo (TAI) | Low | A1B1C0 | + (subpial, pericontusional) | - | Mild | Subacute combined degeneration of spinal cord |
| 4 | 1412 | + (targetoid lesion with hemosiderin in basis pontis; linear lucencies in parietal parasagittal WM) | Linear rusty discoloration (HTT), SFG | "Suspicious" | A0B1C0 (PART) | - | - | Mild | Cavernoma of pons (incidental) |
| 5 | 1200 | + (hemosiderin and volume loss, frontal; torn septum; WM susceptibilities frontal; hydrocephalus ex vacuo; periventricular gray matter nodules) | SDH; contusions; HTTs, thin CC, fenestrated septum (TAI) | Low | A0B2C0 (PART) | + (subpial, pericontusional) | Mild | - | Periventricular nodular heterotopias (incidental) |
| 6 | 1430 | - | Linear rusty discolorations (HTTs), parasagittal WM | Low | AxB1Cx | - | - | Mild | Limited gross examination (received partially cut); focal acute SAH (acute GSW to head) |

**Legend:** AD-NC: Alzheimer Disease neuropathologic change; ARTAG: Aging-related tau astroglipathy; CC: Corpus callosum; CTE-NC: Chronic traumatic encephalopathy neuropathologic change; FTLD: Frontotemporal Lobar Dementia; GSW: Gunshot wound; HTT: Hemorrhagic tissue tears; LBD: Lewy Body Disease; PART: Primary age-related tauopathy; SAH: Subarachnoid hemorrhage; SFG: Superior frontal gyrus; TAI: Traumatic axonal injury; x: missing data; WM: White matter; \* expected = 1200-1500g

Of the 6 cases with CTE-NC, 5 had clear evidence of significant exposure to RHI throughout their lifetimes. Specifically, 2 cases played American football through the college level (for a total exposure of 16 years each; Case 4 & 6) and 1 played professional American football (for 17+ years of exposure; Case 1). One was a boxer (for an unknown period of time) and also sustained an isolated complicated mild TBI (cmTBI; defined as a blow to the head resulting in a period of altered mental status ≤24 hours and/or unconsciousness ≤30 minutes,^33^ but with intracranial findings detected on acute neuroimaging)^34,35^ 2 years prior to death (Case 2). One case served in the military (for 6 years; participated in combat training for 4 years, was deployed for a total of 4 years) and also sustained an isolated cmTBI 2 years prior to death (Case 5). Notably, one case had no known history of RHI but sustained 2 isolated moderate-severe TBIs (msTBI) 30 and 3 years prior to death (Case 3).

### Ex vivo neuroimaging

Four of the 6 cases demonstrating CTE-NC underwent ex vivo neuroimaging; the other 2 were either partially artifactually disrupted or previously sectioned, precluding such examination (Table 2). All 4 imaged specimens had diverse abnormalities detected, including white matter (WM) signal changes and/or volume loss in all 4 (100%), accompanying signal changes of hemosiderin deposition in superficial cortices (in 2 cases [50%]); linear WM signal abnormalities suggesting old hemorrhagic tissue tears (markers of torsional [traumatic axonal] injury) (in 2 cases [50%]); and torn septum pellucidum (in 3 cases [75%]). Incidental findings included congenital periventricular nodules of gray matter signal intensity (in 1 case) and a congenital targetoid vascular lesion with hemosiderin signal in the basis pontis (in 1 case) (not shown). All of these findings guided histologic sampling, allowing co-registration and definitive diagnosis of the underlying tissue substrate (Table 2; Figures 1 and 4).

**Figure 1.**
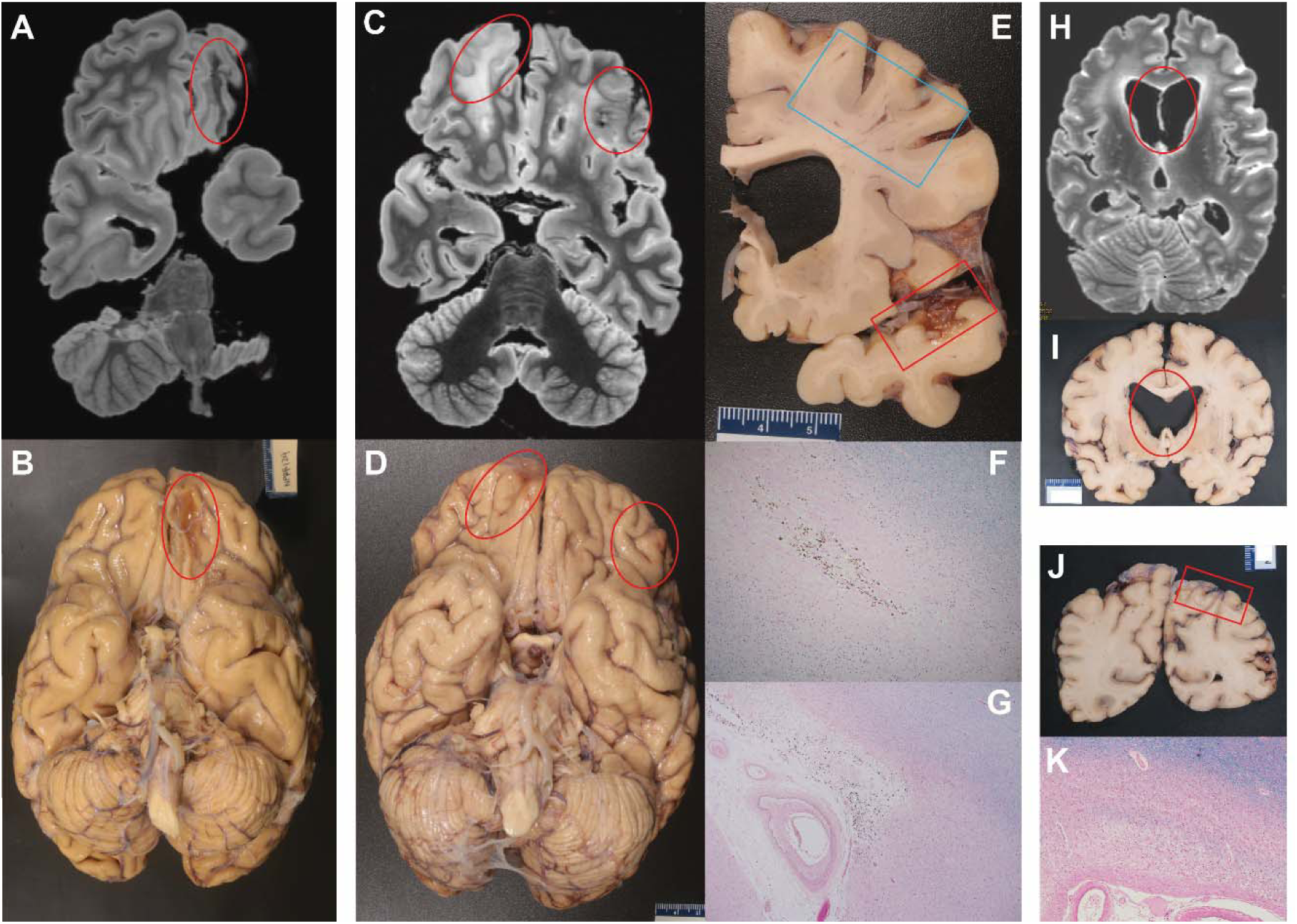
Remote neurotrauma in cases with CTE-NC Case 3 (A, B) with diminished signal on 3T FLAIR-based ex vivo neuroimaging, likely reflecting hemosiderin and corresponding old contusion on gross pathology (red ovals). Case 5 (C, D) also showing hemosiderin signal on 3T FLAIR-based ex vivo neuroimaging indicating sites of prior hemorrhage in old contusions (red ovals). Case 5 (E-G) on cut section with linear rusty discoloration in frontal subcortical WM (blue box) and old temporal contusion (red box); note ventricular expansion indicative of WM volume loss. (F) LHE tissue section of blue box in (E), confirming old HTT as pale scar with brown hemosiderin granules. (G) LHE section of red box in (E) of temporal cortical and subcortical white matter loss with iron deposition (old contusion). Case 2 (H, I) with torn septum pellucidum visualized on ex vivo neuroimaging, confirmed on gross pathology (red ovals); WM volume loss and secondary ventricular expansion are conspicuous; (J, K) Parieto-occipital cortices with “watershed” infarcts (red box), visible on LHE stain as old ischemia having “moth-eaten” appearance of middle layers of cortical ribbon with neuronal loss, macrophages and gliosis. HTT: hemorrhagic tissue tear; LHE: Luxol-fast blue/hematoxylin and eosin stain; WM: white matter

**Figure 2.**
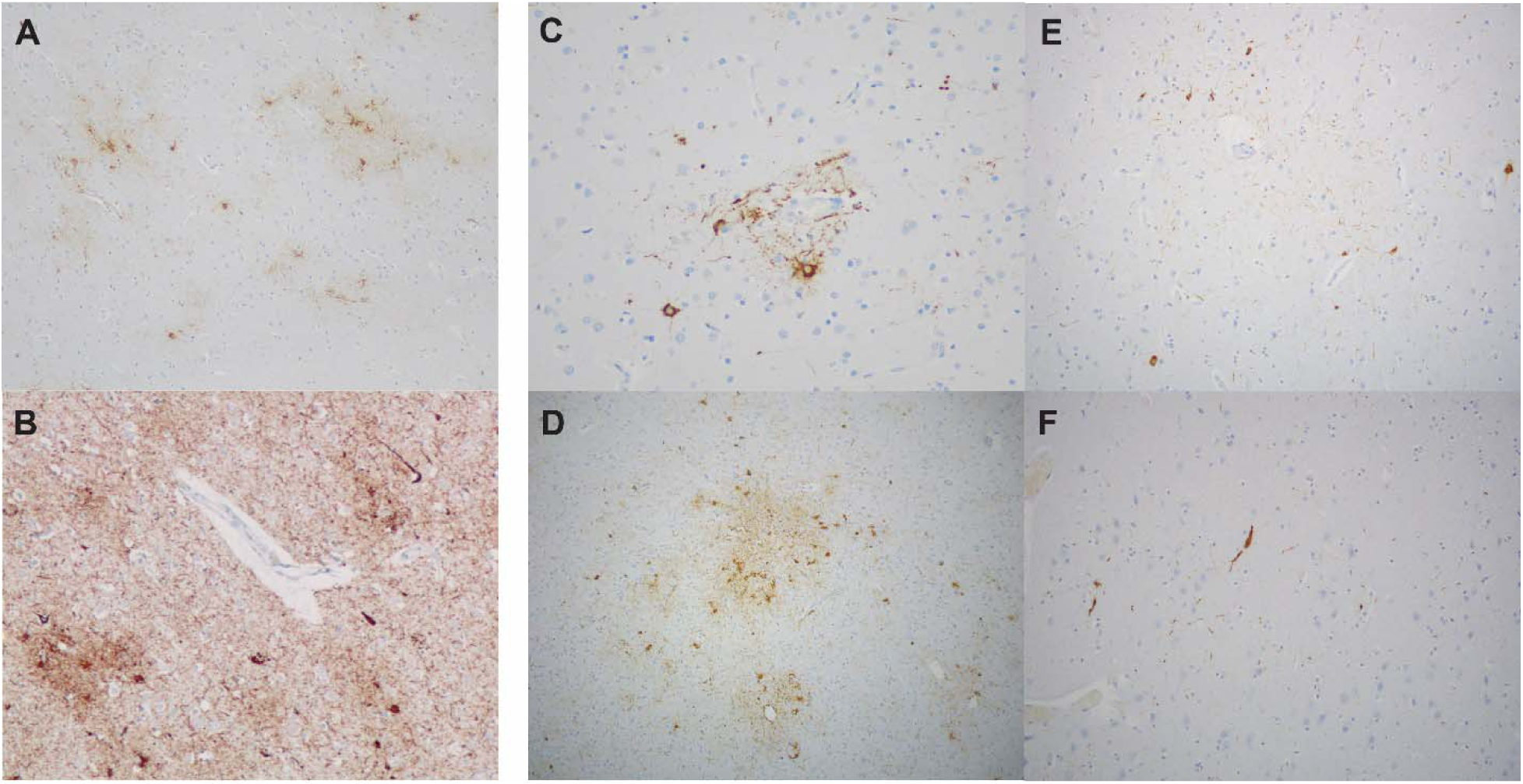
Neocortical phosphorylated tau pathology in the CTE-NC cases Case 1 (A) and Case 2 (B) with high CTE-NC, visible as aggregates of neuronal and astroglial tau in a perivascular distribution in neocortex; subcortical neuronal tau deposition was also significant (not shown). Cases 3, 5, and 6 (C-E) with lesser but distinct perivascular neocortical tau deposition indicative of low CTE-NC. Case 4 (F) with more limited, vaguely perivascular tau deposition considered “suspicious” for CTE-NC. CTE-NC: chronic traumatic encephalopathy-neuropathologic change

### Macroscopic neuropathologic findings

On macroscopic examination, past evidence of remote neurotrauma was evident in 5 of 6 cases (Table 2; Figure 1). Two cases had old subdural hematomas (i.e., neomembranes) (not shown). The ex vivo MRI lesions corresponded macroscopically to old cortical contusions in the 2 cases with hemosiderin signal (Cases 3 and 5) (Table 2; Figure 1); absent or fenestrated septum pellucidum as found in 3 (Cases 2, 3, and 5) (Table 2; Figure 1); old WM hemorrhagic tissue tears as suspected in the finding of linear lucencies in parasagittal white matter in 2 cases (Cases 3 and 4), also found in Case 5 and Case 6 (the latter without ex vivo imaging) (Figures 1 and 4). Diffuse mild or moderate white matter atrophy was confirmed in 3, 2 also with thinning of the corpus callosum and secondary ventricular dilation as noted on ex vivo MRI.

The ex vivo MRI notations of a circumscribed hemorrhagic nodule in the basis pontis of Case 4 (not shown) and periventricular nodular heterotopia in Case 5 (not shown) were documented as incidental (non-trauma-related) findings on gross examination.

### Neurohistologic findings

#### Tau and other proteinopathies

On histology, 2 cases showed changes of “high” CTE-NC, while 3 cases were assigned a “low” CTE-NC burden; one had limited tau deposition and was considered “suspicious” for CTE-NC (Figure 2).

“High” CTE-NC

Case 1 (7^th^ decade, football for 17 years) showed a high density of scattered phosphorylated tau-labeled neuronal pretangles, tufted astrocytes, and thorny astrocytes as well as glial cytoplasmic inclusions, often in a patchy, irregular pattern throughout most cortical areas, of greatest severity at the depths of sulci in the frontal and temporal lobes (Figure 2). The basal ganglia and brainstem demonstrated low densities of neurofibrillary tau, and the hippocampus and entorhinal cortex showed neurofibrillary pathology with predominance in sectors CA2 and CA4 (not shown). Aging-related tau astrogliopathy (ARTAG) was noted in subpial regions (Figure 3). Alzheimer’s disease neuropathologic change (ADNC) consisted only of tau (A0B1C0), possibly representing very early changes of primary aging-related tauopathy (PART).

**Figure 3.**
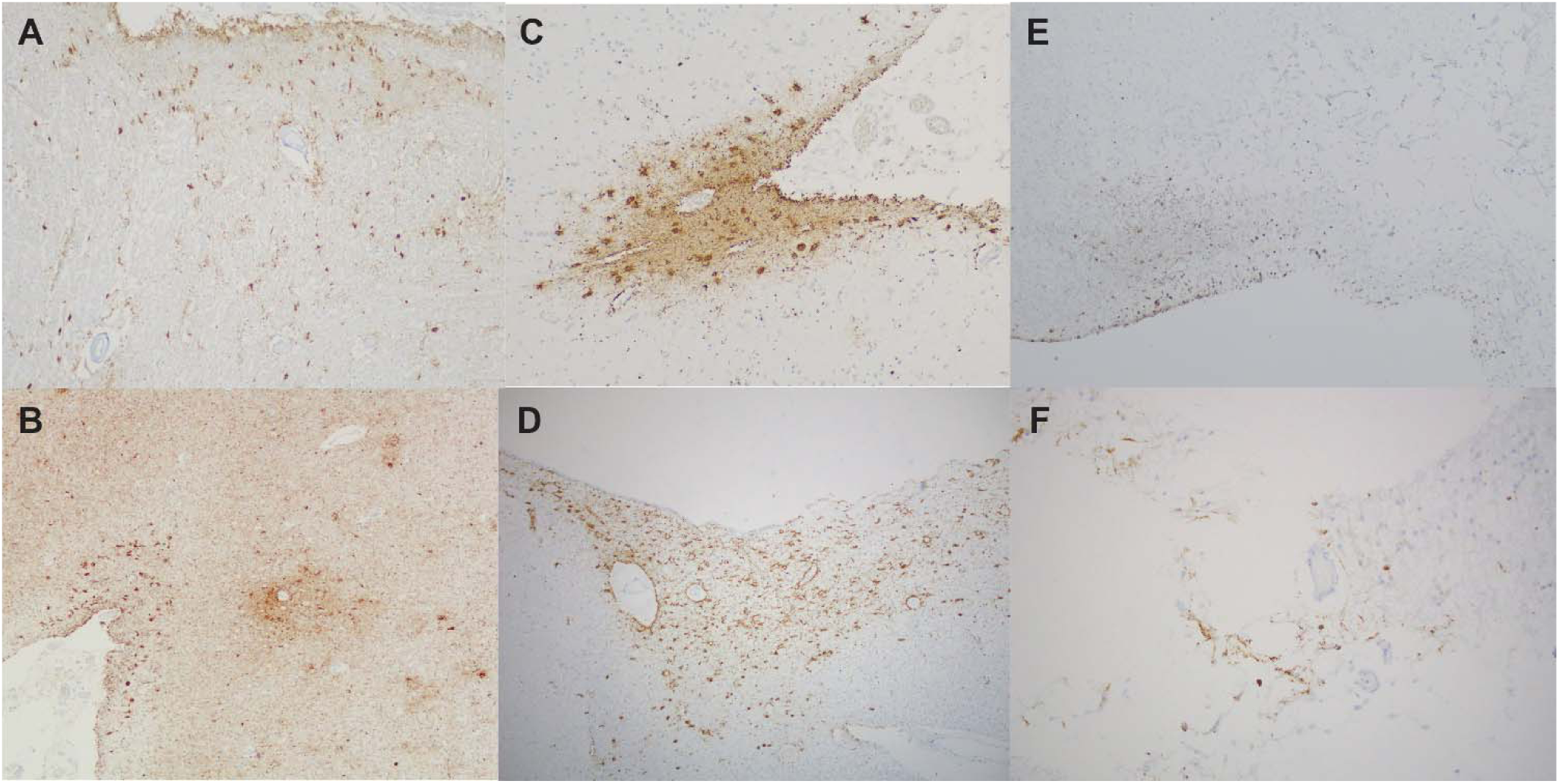
Additional phosphorylated tau deposition in the CTE-NC cases Case 1 (A), Case 2 (B), and Case 3 (C) with subpial aging-related tau astrogliopathy (ARTAG). Case 5 (D) with subependymal and perivascular white matter ARTAG. Cases 3 and 5 (E, F) contusions with cavitation and perilesional tau.

**Figure 4.**
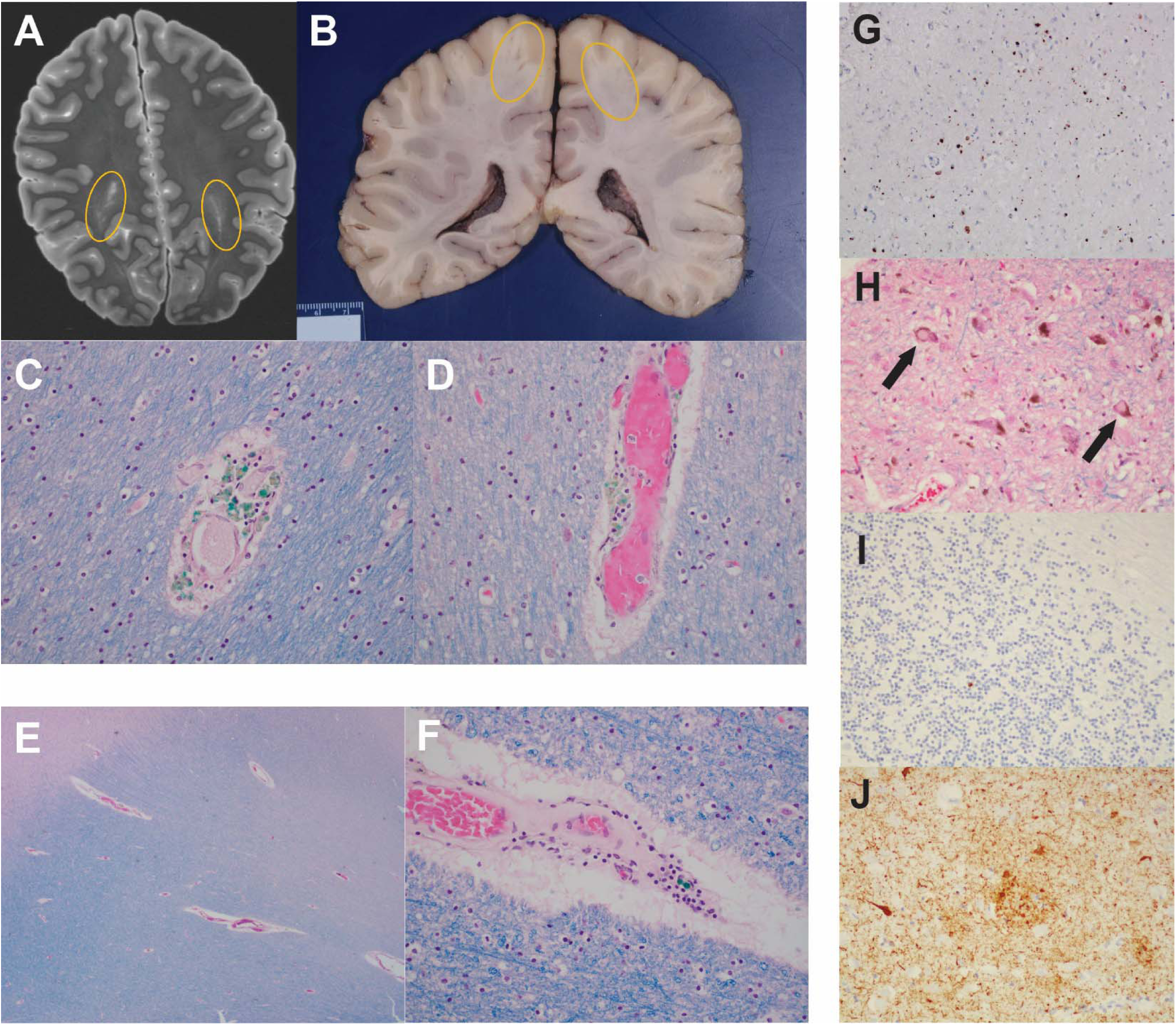
Co-occurring pathology in cases with CTE-NC Case 4 (A, B) Parasagittal linear signal on 3T FLAIR-based ex vivo neuroimaging, corresponding to fine rusty streaks in the subcortical white matter (yellow ovals); LHE sections (C, D) demonstrating these to be regions of widened perivascular spaces containing brown pigment- and green myelin debris-laden macrophages, as well as perivascular myelin pallor, consistent with prior torsional vascular injury. Case 6 (E, F) with similar changes, but thicker collagenized vascular wall attributed to hypertensive small vessel (arteriolosclerotic) disease. Case 2 (G-J) with TDP-43 immunostaining of hippocampal neurons; Lewy Body Disease (arrows) in the brainstem, also seen in limbic system (not shown); (I) rare p62- (ubiquitin) immunopositive inclusions in the cerebellar granular layer, also in dentate gyrus (not shown); (J) Bielchowsky silver preparation revealing neuritic plaques and neurofibrillary tangles representative of high AD-NC. AD-NC: Alzheimer Disease-neuropathologic change; LHE: Luxol-fast blue/hematoxylin and eosin stain;

In Case 2 (7^th^ decade, boxing for unknown duration + cmTBI), the phosphorylated tau immunostains of hippocampi and neocortex were positive for frequent neurofibrillary tangles, variably dense neuropil threads and fuzzy diffuse granular cortical staining (Figure 2). Often, neurofibrillary tangles and thorny astrocytes occupied a perivascular distribution at the depths of sulci; in addition, there was a predominance of phosphorylated-tau immunostaining in the CA2 and CA4 hippocampal subregions (not shown). ARTAG pathology was also observed, primarily in subpial locations (Figure 3).

Amyloid-beta immunostains and Bielschowsky silver preparations revealed high AD-NC (A3B3C2). Limbic-stage Lewy Body Disease was confirmed on alpha-synuclein immunostaining (Figure 4). TDP-43-immunopositive neurons were conspicuous in the frontal and temporal cortices, primarily in the upper and lower layers as well as in the hippocampal CA1 subregion, entorhinal cortex and dentate gyrus granule cell neurons (Figure 4). Rare p62 (ubiquitin) immunopositive inclusions were found in the dentate gyrus and cerebellar granular cell neurons (Figure 4). Thus, this case represents a “polyproteinopathy”.

#### “Low” CTE-NC

Case 3 (6^th^ decade, 2 msTBIs), Case 5 (7^th^ decade, army for 6 years + cmTBI) and Case 6 (4^th^ decade, football x 16 years) showed predominantly neocortical, mild tau deposition that was focally perivascular or at the depths of cortical sulci (Figure 2). Case 4 was “suspicious” for CTE-NC on the basis of p-tau in superficial frontal cortex (Figure 2), as well as threads in the hippocampus (not shown). Phosphorylated-tau immunostaining also revealed ARTAG in Cases 3 and 5 (Figure 3). Phosphorylated tau was conspicuously associated with old contusions in Cases 3 and 5 (Figure 3), alongside frontotemporal cavitation and hemosiderosis as detected by ex vivo MRI and macroscopic inspection.

### Torsional (traumatic axonal and/or vascular) injuries

In addition to the lesions of the septum pellucidum noted on ex vivo MRI and confirmed macroscopically in 3 of the 6 cases, other evidence of torsional injury to the brain comprised linear lucencies in the parasagittal white matter (Cases 3 and 4) corresponding to old hemorrhagic tissue tears/perivascular hemosiderin, also noted pathologically in Cases 5 and 6 (Figure 1). White matter volume loss, including callosal thinning, and secondary ventricular expansion (Figure 1), particularly affecting Cases 3 and 5, are supportive evidence of significant traumatic axonal/vascular injury.

### Cerebrovascular disease

Case 1 had moderate athero- and arteriolosclerosis. In Cases 2, 3, and 4, periventricular signal changes on ex vivo MRI corresponded to mild white matter microvascular disease documented histologically, also found in Case 6 (Figure 4). Case 2 was notable for old “watershed” infarcts in bilateral parieto-occipital regions (Figure 4), detected as cortical hyperintensity on ex vivo MRI (not shown). Slight atherosclerosis was documented in Case 5. No cases had microinfarcts or cerebral amyloid angiopathy.

## DISCUSSION

Among 44 autopsied brain donors in the Late Effects of TBI (LETBI) Study brain donor program, 6 (13.6%) had autopsy-confirmed CTE-NC (1 of which had very limited burden, only considered “suspicious” for CTE-NC). Two of these 6 were categorized as “high” CTE-NC and 3 as “low” CTE-NC. Only one of the cases with any CTE-NC had no evidence of RHI; this was a single case with “low” CTE-NC in an individual who sustained 2 isolated TBIs but no documented history of RHI.

### Significance of isolated versus repetitive head injury

Reports of CTE-NC (as defined by the second consensus paper) arising in the context of isolated TBI remains relatively rare, with findings from case reports and brain bank series alike yielding limited supportive evidence. Ling et al reported CTE-like p-tau immunostaining patterns in 32/268 degenerative disease brain bank cases screened for this finding: these 32 cases included 30 known to have had TBI with or without loss of consciousness. Eleven participated in contact sport and 6 were military veterans. All but 4 had neurodegenerative diseases known to be associated with frequent falls. Of note, this study also found “low” CTE in 12% of controls.^36^ Tau pathology (notably not pathognomonic for CTE-NC) has been reported in only occasional survivors of single TBI.^37,38^ Rather, several studies report that non-CTE tau deposition following single TBI may co-occur with multiple neurodegenerative pathways, inducing Alzheimer’s Disease, Lewy Body Disease and TDP-43 proteinopathy.^36,37,39,40^ We speculate that the polyproteinopathy noted in one of our cases may be such an example, and further discuss other neuropathologies found among our cohort.

### Co-existence of other neuropathologic features

#### Tau deposition in ARTAG and around contusions

The pathogenesis and significance of ARTAG has been debated since its initial recognition among older individuals in neurodegenerative brain banks, often but not always accompanied by AD-NC or other types of neurodegeneration.^32,41^ Its more recent detection among younger people with a variety of disorders, including intimate partner violence in women in their 3rd to 5th decades,^21^ has prompted consideration of modifying the term to “tau astrogliopathy” (TAG), however, this has not been adopted formally.

Regarding “perilesional” ARTAG (a subtype specified but not illustrated in Kovacs et al., 2016)^32^, few studies have documented tau deposition adjacent to old contusions, as we illustrate here in both of our 2 cases with old contusions. Notable among these is the study by Noy et al., who found tau (CTE-like) adjacent to old contusions in 1 of 5 cases with both low-stage CTE-NC and old contusions;^42^ the same laboratory offered a follow-up study in which pericontusional ARTAG was reported in 2 of 6 cases with CTE-NC and old contusions.^12^ In unpublished observations in a busy forensic practice over 9 years by one of us (RDF), perilesional tau deposition was noted not infrequently adjacent to old contusions, gunshot-wound tracks, and in a 13-year-old survivor of abusive head trauma in infancy, again not necessarily an “aging-related” phenomenon.

Aside from impact-related contusions, CTE-NC has also been identified in areas of iatrogenic primary axonal injury, such as leucotomy sites in 4 of 5 long-term survivors of schizophrenia patients undergoing that procedure (now of historical interest), as well as adjacent to arteriovenous malformation, old infarct, and glioblastoma, accompanied by ARTAG-like tau.^43,44^ Other authors have evaluated TBI series and found no tau deposition adjacent to old traumatic injuries, beyond that seen in age-similar subjects.^45^

The assumed mechanism of generation and accumulation of p-tau is based on the observation that microtubule-associated protein tau is released from the axons following stretching or tearing forces, and may undergo misfolding, abnormal phosphorylation, and deposition within and outside of cells and their processes. Related phenomena have been proposed to contribute, namely, endoplasmic reticulum stress and blood-brain barrier disruption,^8,46–51^ which of course are not unique to mechanical injury but may follow a wide range of cellular disruptions. The potential significance of alcohol and/or substance use disorder, independent of their associated propensity for trauma, may themselves be risk factors for tau accumulation.^12,42,52,53^

### Non-tau-related mechanical effects of TBI

Substantial direct (contusional) impact, leading to cortical loss and often extensive subcortical white matter degeneration, as well as torsional (rotational, “whiplash”-type) injuries resulting in hemorrhagic tissue tears, both result in substantial volume loss, as reflected by atrophy, lower-than-expected brain weight, and variable secondary ventricular expansion (hydrocephalus ex vacuo). As has been shown in our prior work in victims of intimate partner violence (IPV) and by the work of Johnson et al, markers of traumatic axonal injury may persist for years and indicate ongoing reactive/degenerative processes involving neuroinflammation (beyond the scope of this article).^21,54^ Thus, the clinical meaning of CTE-NC when accompanied by sequelae of major trauma may be limited if not inconsequential.

### Microvascular injury

Aside from direct cortical impact trauma resulting in frank contusions (by definition causing localized vascular compression, disruption of the blood-brain barrier, ischemia and hemorrhage, as well as downstream neuroinflammatory sequelae,^55,56^ often persisting for years), subconcussive RHI injury to cortical gray matter has focused particularly on microvessels at sulcal depths, where CTE-NC lesions predominate. Quantitation of vascular branch density and fraction volume in cortical samples demonstrated increases in both measures among CTE cases versus controls.^57^ Work by the same group also found increased levels of vascular-injury associated markers, such as intercellular adhesion molecule 1, vascular cell adhesion molecule 1, and C-reactive protein, and association with increased microglial density and tau pathology, particularly at sulcal depths.^58^ Clinical imaging studies of white matter hyperintensities in former American football players indicate a greater burden of findings in older men compared to age-similar men not exposed to contact sports, and also to younger men previously exposed.^59^ In studies by the same group, the neuropathologic substrate (primarily defined in semiquantitative terms as white matter rarefaction and arteriolosclerosis) was correlated with antemortem imaging findings: both were significantly associated with increased odds of dementia, independent of tau, in subjects >40 years old.^60^ The contribution of old traumatic axonal injury was not analyzed per se.

In the current study, we focused on white matter microvascular changes, highlighted on ex vivo imaging in our cohort, and confirmed in a point-by-point correlative manner with the underlying tissue substrate, as previously reported by us in women victims of intimate partner violence (IPV) with TBI and nonfatal strangulation.^21^ Based on that study and the current one, we propose a means of histopathological discrimination between those vessels with characteristic features of arteriolosclerosis versus those that may have trauma-induced alterations: briefly, both may have similar imaging characteristics, reflecting widened perivascular spaces, perivascular rarefaction and myelin pallor, and deposition of non-heme iron pigment, although hypertensive “small vessel” disease is defined histologically by collagenous wall thickening. This important diagnostic feature was notably absent in some imaging-guided sections, and instead suggested primary traumatic blood-brain barrier interruption and, in a subset, microbleeds derived from torsional injury. In the IPV cohort, we further evaluated brain sections with immunostains for fibrinogen (blood-brain barrier leakiness), microglial/macrophage (inflammatory) markers, and iron deposition (resulting from microbleeds); these additional studies are currently in progress for all of our 44 postmortem brains in the LETBI study.

### Limitations of this study

The accrual of postmortem human brain tissue donations has many well-recognized limitations, including selection bias (for educated, motivated individuals and their families with access to healthcare, and for those with more severe trauma and post-traumatic sequelae). Clinical and autopsy catchments are not always hospital-based, such that trauma-related deaths may fall under local medicolegal jurisdictions, and may be harder to obtain in a timely manner.

We acknowledge the difficulty in ascertainment of prior RHI exposure among autopsy cases for whom medical record access and/or family recollection, which may be the case in many brain banks whether collecting sport, military, or neurodegenerative specimens. We therefore emphasize the advantage of the LETBI study, which uses all available information for all types of trauma exposure ascertainment – including prospectively collected self-report data, detailed clinical records, police reports, autopsy reports, and publicly available internet records – to optimize accuracy of lifetime head trauma exposure characterization.

## Conclusions

In a consecutive case series of 44 autopsy brains from the Late Effects of Traumatic Brain Injury donor cohort, we report 6 cases with neuropathologic features of CTE, including 1 with a history of 2 isolated TBIs but without RHI. In concordance with other studies,^53^ our data suggest that RHI may not be an absolute requirement for the development of progressive neurodegeneration, specifically CTE-NC. Moreover, we raise concern regarding the clinical significance of “low” CTE-NC burden in light of other contusional and torsional mechanical injuries and their long-term sequelae, including microvascular disease, as well as co-morbid proteinopathies. The finding of ARTAG in individuals in the 3^rd^ and 5^th^ decades (as noted in previous studies in IPV from our group)^21^ further prompts reconsideration of whether the modifying term “aging-related” is appropriate.

## Data Availability

All data produced in the present study are available upon reasonable request to the authors

## ACKNOWLEDGEMENTS

National Institutes of Health/National Institute on Neurological Disorders and Stroke: RF1NS115268, RF1NS128961

National Institutes of Health/National Institute on Neurological Disorders and Stroke/National Institute of Child Health and Development: U01NS086625

NIH Research Training in the Neuroscience of Aging: T32AG049688

We are grateful to Dr. John F. Crary, Dr. Jamie Walker, Dr. Etty Cortes, and Dr. Adam Goldstein, and other members of the Neuropathology Brain Bank at Mount Sinai, for their work and contributions to this project.

